# Timing is everything: the relationship between COVID outcomes and the date at which mask mandates are relaxed

**DOI:** 10.1101/2021.03.31.21254646

**Authors:** Affan Shoukat, Alison P. Galvani, Meagan C. Fitzpatrick

## Abstract

**Importance:** Several states including Texas and Mississippi have lifted their mask mandates, sparking concerns that this policy change could lead to a surge in cases and hospitalizations.

**Objective:** To estimate the increase in incidence, hospitalizations, and deaths in Texas and Mississippi following the removal of mask mandates, and to evaluate the relative reduction of these outcomes if policy change is delayed by 90 days.

**Design, Setting, and Participants:** This study uses an age-stratified compartmental model parameterized to incidence data in Texas and Mississippi to simulate increased transmission following policy change in March or June 2021, and to estimate the resulting number of incidence, hospitalizations, and deaths.

**Main Outcomes and Measures:** The increase in incidence, hospitalizations, and deaths if mask mandates are lifted on March 14 compared to lifting on June 12.

**Results:** If transmission is increased by 67% when mask mandates are lifted, we projected 11.39 (CrI: 11.22 - 11.55) million infections, 170,909 (CrI: 167,454 - 174,379) hospitalizations, and 5647 (5511 - 5804) deaths (Figure 1) in Texas from March 14 through the end of 2021. Delaying NPI lift until June reduces the average number of infections, hospitalizations, and deaths by 36%, 65%, and 62%, respectively. Proportionate differences were similar for the state of Mississippi. Peak hospitalization rates would be reduced by 79% and 63% in Texas and Mississippi, respectively.

**Conclusions and Relevance:** Removal of mask mandates in March 2021 is premature. Delaying this policy change until June 2021, when a larger fraction of the population has been vaccinated, will avert more than half of the expected COVID-19 hospitalizations and deaths, and avoid an otherwise likely strain on healthcare capacity.

## Main Text

In an extraordinary achievement, multiple vaccines against SARS-CoV-2 have been developed within a year of pathogen identification. As of early March 2021, three vaccines were approved for emergency use by the United States Food and Drug Administration. Vaccine rollout began on December 14, 2020 and the US government plans to provide all adults with access to vaccines by the end of May 2021.^1^

**Figure 1:**
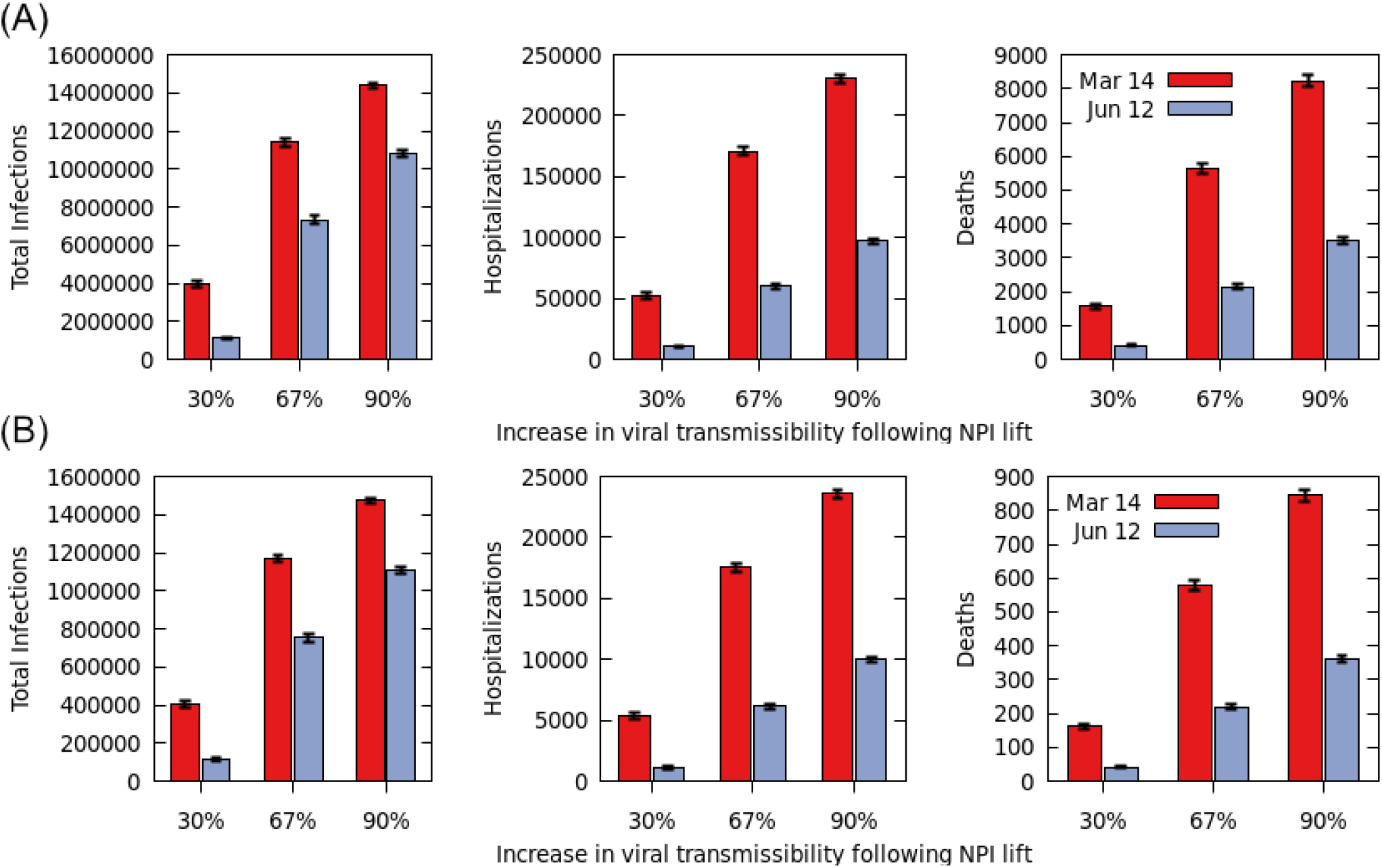
Cumulative number of infections, hospitalizations, and deaths for (A) Texas and (B) Mississippi through 2021 if NPI were lifted on March 14 (red) or June 12 (blue).

Prior to the initiation of vaccine rollout, the United States experienced its largest wave of SARS-CoV-2 infections, hospitalizations, and deaths. In light of recently declining incidence and rising vaccination coverage, political leaders in several states, including Texas and Mississippi, have lifted non-pharmaceutical interventions (NPI), such as mask mandates.^1^ However, the burden still exceeds early fall levels and the spread of highly transmissible variants may reverse the declining trend. Given the rapidly shifting epidemiological landscape of SARS-CoV-2, evidence to guide optimal timing for relaxation of NPI has been scarce.

## Methods

To project the impact of NPI relaxation timing on incidence, hospitalizations and deaths, we developed an age-stratified SARS-CoV-2 transmission model. We calibrated the model to incidence data in Texas and Mississippi from December 14 to March 3, 2021, incorporating state-specific demography and pre-existing immunity (Appendix). Social mixing patterns were derived from empirical studies conducted both prior to and during 2020.^2,3^ Upon transmission, individuals enter a latent stage from which an age-specific proportion proceeds to an asymptomatic infectious stage.^4^ The remainder transition to a highly infectious presymptomatic stage, followed by symptom onset, incorporating age-specific probabilities of clinical severity. Infected individuals who recover are fully protected against subsequent infection. A two-dose vaccination campaign was implemented according to a temporal distribution strategy that replicated the age-specific allocation in each state as of March 3.^5^ For allocation after that date, vaccines were distributed according to those age-specific ratios until coverage reached age-specific acceptance rates for COVID-19 vaccines based on survey data.^6^ Vaccinated individuals received two doses 21 days apart, with efficacies against symptomatic disease of 66% and 94% for first and second doses, respectively, and against infection of 46% and 92%, respectively.^7,8^ Model parameters were sampled from their statistical distributions, and clinical outcomes were averaged over stochastic 1000 realizations from March 14 through the end of 2021.

We simulated the effect of lifting NPI as an increase in the transmissibility of SARS-CoV-2. To represent the lifting of mask mandates as the base case, we raised the probability of infection given contact between a susceptible and infectious individual by 67% above its calibrated value, consistent with empirical assessment of the protection afforded by masks.^9^

## Results

With the lifting of NPI on March 14, we projected 11.39 (CrI: 11.22 - 11.55) million infections, 170,909 (CrI: 167,454 - 174,379) hospitalizations, and 5647 (5511 - 5804) deaths (Figure 1) in Texas from March 14 through the end of 2021 (Figure 1). Delaying the lift by 90 more days would markedly improve health outcomes. For instance, this delay would result in 7.34 (7.13M - 7.55M) infections, 60,122 (58,299 - 61,883) hospitalizations, and 2145 (2071- 2210) deaths (Figure 1). For Mississippi, an early lift results in 1.17 (1.15 - 1.19) million infections, 17,541 (17,187 - 17,897) hospitalizations, and 580 (566 - 596) deaths. By contrast, delayed lift results in 753,807 (731,801 - 774,822) infections, 6171 (5984 - 6351) hospitalizations, and 220 (213 - 227) deaths.

Compared to early lift, delaying NPI lift until June reduces the average infections, hospitalizations, and deaths by 36%, 65%, and 62%, respectively, in both Texas and Mississippi. Delay also alleviates pressure on the healthcare system, reducing peak hospitalization by 79% and 63% in Texas and Mississippi, respectively (Figure 2). Across both states, we project that a cumulative 3682 deaths could be averted if lifting NPI were delayed by 90 days.

**Figure 2.**
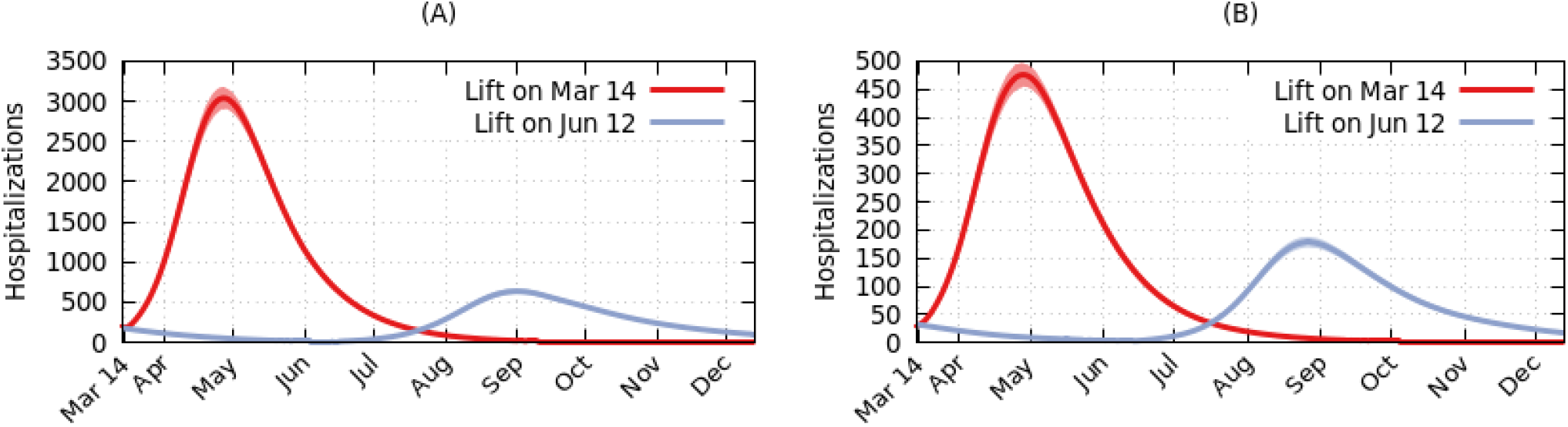
Daily new hospital admissions when NPI were lifted on March 14 (red) or June 12 (blue) in (A) Texas and (B) Mississippi with 67% increased transmissibility.

Our scenario analyses indicated that with a larger increase in viral transmissibility after the lifting of NPI, the proportionate impact of a delay declines but the magnitude of the impact is greater (Figure 1, Figure A3, Table A3). For instance, if transmissibility increases by only 30% instead of 67% following NPI lift in Texas, a delay from March to June could avert 41,177 hospitalizations, equivalent to a 79% reduction compared to early NPI lift. If NPI lift results in 90% increased transmissibility for Texas, a delay could avert 132,786 hospitalizations which is 57% of the expectation for an earlier lift.

## Discussion

The incidence of COVID-19 cases has declined across much of the United States since mid-January, principally due to ongoing NPI such as mask mandates, social distancing measures, and travel restrictions. This declining trend would be expected to continue as vaccination is rolled out across the country. Political leaders in several states have recently interpreted the downward trend in cases as a reason to lift NPI in March 2021, despite the fact that only 10% of the US population had received a full vaccine regimen as of March 9.^5^ Our results indicate that these decisions are premature. In both Texas and Mississippi, a removal of mask mandates in March will lead to substantially higher epidemic peaks and more cumulative infections compared to a delay of even three months.

If the objective of lifting NPI is a return to normalcy and economic productivity, doing so in March is likely to be counterproductive. For instance, the precipitous rise in hospitalizations that results from suspending mask mandates for Texas in March would exceed that state’s winter surge,^10^ likely necessitating even stricter NPI in order to return to current COVID-19 levels. By contrast, maintaining mandates through June would avert this hospitalization crisis.

Based on studies of the protection conferred by masks, in our base case we specified that individual-level risk per infectious contact would rise by 67% under policies which lift NPI. However, the change in transmission dynamics will depend on several factors, including the responsiveness of local jurisdictions and businesses to state-level policy change. For instance, several Texas cities including the capital announced that local mask mandates will replace those discontinued by the state.^11^ Due to the nonlinear relationship between transmissibility and outbreak size, local and individual actions to maintain mask use can have an outsized protective effect. Conversely, simultaneous reversal of additional NPI, such as density restrictions, may amplify the increase in transmissibility.

In two respects, our results conservatively underestimate the number of hospitalizations and deaths which could be prevented by delaying NPI lift until June. First, several variants of SARS-CoV-2 that appear to be more transmissible than the currently dominating strain have emerged in the United States. In particular, the B.1.1.7 variant is estimated to be 43-90% more transmissible than the originally dominant US variant.^12^ Alarmingly, vaccine protection against some variants may be compromised.^13,14^ Nevertheless, if even partial vaccine protection exists,^15^ variant spread will be most rapid if NPI are lifted prior to widespread vaccination coverage rather than later. Second, SARS-CoV-2 transmission may be dampened during the North American summer, relative to early spring, consistent with the seasonality observed in 2020. If so, then viral transmissibility will likely be lower following NPI lift in June compared to March, and the benefits of delay are expanded beyond those we project here.

Our results suggest that, synergistic with a vaccination campaign, continuing public health measures such as proper masking and maintaining social distancing are critical for curbing SARS-CoV-2 transmission. Hasty abandonment of these fundamental control measures prior to widespread vaccination coverage will likely result in both the need for even stricter measures, as well as loss of life.

## Data Availability

Not Applicable.

## Acknowledgments

The authors are grateful for funding from the Notsew Orm Sands Foundation, National Science Foundation (Expeditions 1918784; RAPID-2027755), Centers for Infectious Disease Control and Prevention (201903979), and National Institutes of Health (K01AI141576; 1R01AI151176).

## Notes

### Competing Interest Statement

The authors have declared no competing interest.

