## Appendix for "Timing is everything: the relationship between COVID outcomes and the date at which mask mandates are relaxed"

#### 1 THE MODEL

We modelled the transmission of SARS–CoV-2 using an age-structured compartmental model. We included standard SEIR compartments as well as additional compartments to reflect asymptomatic infections, presymptomatic transmission, and vaccination. See Table A1 for a full description of the compartments and Figure A1 for a model schematic. We assume that the population is large and closed, and therefore do not consider births, deaths, and migration. The model is given by the following system of nonlinear differential equations:

$$
\begin{aligned}
S_a' &= -S_a\mathscr{F}_a - \lambda_{s,a}\\
V_{1,a}' &= \lambda_{s,a} - \lambda_{v,a} - (1-\varepsilon_{1,a})V_{1,a}\mathscr{F}_a\\
V_{2,a}' &= \lambda_{v,a} - (1-\varepsilon_{2,a})V_{2,a}\mathscr{F}_a\\
E_a' &= S_a\mathscr{F}_a - \sigma E_a\\
F_{1,a}' &= (1-\varepsilon_{1,a})V_{1,a}\mathscr{F}_a - \sigma F_{1,a}\\
F_{2,a}' &= (1-\varepsilon_{2,a})V_{2,a}\mathscr{F}_a - \sigma F_{2,a}\\
A_a' &= p_a\sigma E_a + \rho_{1,a}\sigma F_{1,a} + \rho_{2,a}\sigma F_{2,a} - \eta A_a\\
P_a' &= (1-p_a)\sigma E_a + (1-\rho_{1,a})\sigma F_{1,a} + (1-\rho_{2,a})\sigma F_{2,a} - \theta P_a\\
I_a' &= \theta P_a - h_a\kappa I_a - (1-h_a)(1-f_a)\gamma I_a - (1-h_a)f_a\tau I_a\\
\widetilde{I}_a' &= (1-h_a)f_a\tau I_a - \left(\frac{\tau\gamma}{\tau-\gamma}\right)\widetilde{I}_a\\
X_a' &= h_a\kappa I_a - (1-d_a)\nu X_a - d_a\nu X_a\\
D_a' &= d_a\nu H_a\\
R_a' &= \eta A_a + (1-h_a)(1-f_a)\gamma I_a + \left(\frac{\tau\gamma}{\tau-\gamma}\right)\widetilde{I}_a + (1-d_a)\nu X_a
\end{aligned}
$$

where $\mathscr{F}_a$ is the age-specific force of infection, given by

$$
\mathscr{F}_a = \beta\left(\sum_{j=1}^{5} M_{a,j}\frac{(\alpha A_j + P_j + I_j)}{N_j} + \sum_{j=1}^{5}\tilde{M}_{a,j}\frac{\widetilde{I}_j}{N_j}\right)
$$

The transmission parameter $\beta$ was calibrated to reported incidence in the states of Texas and Mississippi from December 14 to March 3 (Figure A2). Upon transmission, individuals enter a latent stage from which an age-specific proportion proceeds to an asymptomatic infectious stage [1]. The remainder transition to a highly infectious presymptomatic stage, followed by symptom onset, incorporating age-specific probabilities of clinical severity. Infected individuals who recover are fully protected against subsequent infection. Model parameters were sampled from their statistical distributions when available, and clinical outcomes were averaged over 1000 stochastic realizations from March 14 through the end of 2021. Social mixing patterns were derived from empirical studies conducted both prior to and during 2020. In particular, transmission between and within age groups was based on heterogeneous mixing with rates determined by age-specific contact matrices for regular contacts $M$ and during isolation $\tilde{M}$ [2, 3].

$$M = \begin{array}{c} \begin{matrix} 0-4 & 5-19 & 20-49 & 50-64 & 65+ & \textbf{Age} \end{matrix} \\ \begin{bmatrix} 2.34 & 1.88 & 4.31 & 1.14 & 0.55 \\ 0.46 & 10.02 & 4.83 & 0.99 & 0.49 \\ 0.52 & 2.01 & 8.63 & 1.96 & 0.68 \\ 0.27 & 1.23 & 5.48 & 3.07 & 1.21 \\ 0.17 & 0.87 & 3.26 & 1.75 & 1.96 \end{bmatrix} \begin{matrix} 0-4 \\ 5-19 \\ 20-49 \\ 50-64 \\ 65+ \end{matrix} \end{array},$$

and

$$\tilde{M} = \begin{array}{c} \begin{matrix} 0-4 & 5-19 & 20-49 & 50-64 & 65+ & \textbf{Age} \end{matrix} \\ \begin{bmatrix} 0.65 & 0.53 & 1.21 & 0.32 & 0.15 \\ 0.13 & 2.8 & 1.35 & 0.28 & 0.14 \\ 0.15 & 0.56 & 2.41 & 0.55 & 0.19 \\ 0.08 & 0.34 & 1.53 & 0.86 & 0.34 \\ 0.05 & 0.24 & 0.91 & 0.49 & 0.55 \end{bmatrix} \begin{matrix} 0-4 \\ 5-19 \\ 20-49 \\ 50-64 \\ 65+ \end{matrix} \end{array}$$

where, in each matrix, the elements $\{m_{ij} \mid i, j \in (1, \cdots, 5)\}$ denote the average contact rates between age groups $i$ and $j$. For simulating the model, we used a non-standard numerical method to solve the system of equations.

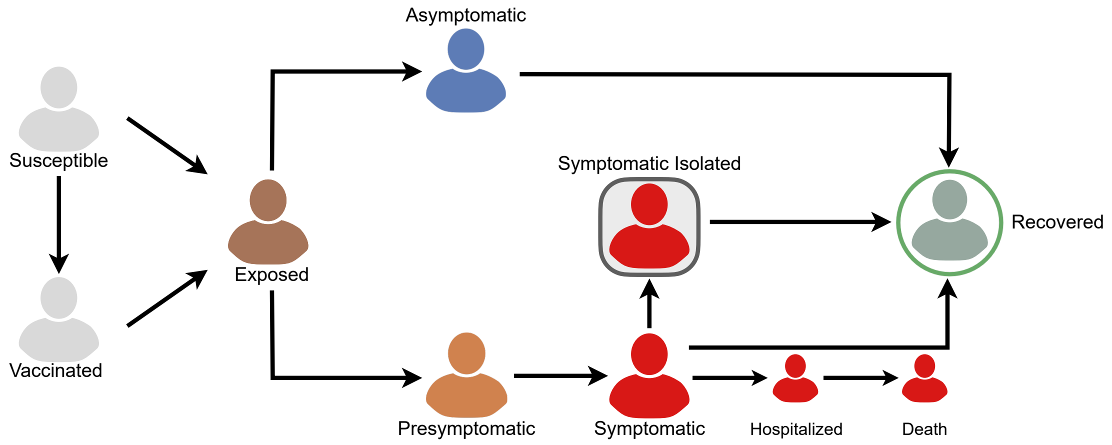


**Figure A1.** Model Schematic

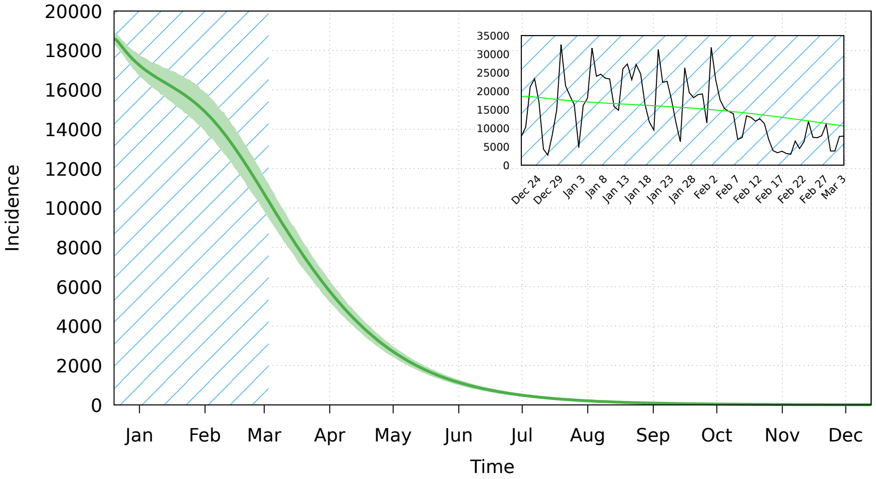


**(a)**

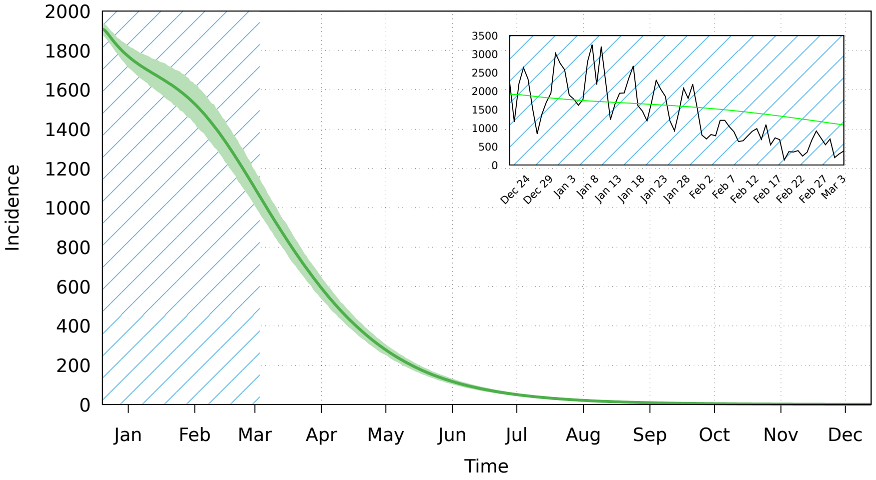


**(b)**

**Figure A2.** Main plots shows a base case scenario which includes vaccination and no lifting of NPI in (a) Texas and (b) Mississippi. Inset plots shows the model fit to reported data from December 14 to March 3.

**Table A1.** Description of the model compartments. The subscript $a$ denotes the age group corresponding to either: 1-4, 5-19, 20-49, 50-65, and 65+.

| Variable | Description |
|---|---|
| $S_a$ | Susceptible |
| $V_{1,a}, V_{2,a}$ | Vaccinated with first or second dose, respectively |
| $E_a$ | Latent |
| $F_{1,a}, F_{2,a}$ | Latent but vaccinated with first or second dose |
| $A_a$ | Asymptomatic |
| $P_a$ | Pre-symptomatic |
| $I_a, \widetilde{I}_a$ | Symptomatic and Isolated Symptomatic |
| $X_a$ | Hospitalized |
| $D_a$ | Death |
| $R_a$ | Recovered |
| $N_a$ | Population size |

**Table A2.** Description of the model parameters and their associated values.

| | **Description** | **Value** | **Source** |
|---|---|---|---|
| $\beta$ | Transmission parameter | Calibrated | |
| $\alpha$ | Relative transmission of asymptomatic individuals | 0.75 | [4] |
| $\lambda_{s,a}, \lambda_{v,a}$ | Daily vaccination rates for 1st and 2nd doses | Calibrated | |
| $p_a$ | % of infected individuals that are asymptomatic | | |
| | ▷ Age group: $0-4$ | 30% | [5] |
| | ▷ Age group: $5-19$ | 37.3% | |
| | ▷ Age group: $20-49$ | 32.8% | |
| | ▷ Age group: $50-64$ | 32.8% | |
| | ▷ Age group: $65+$ | 18.8% | |
| $1/\sigma$ | Latent Period [1] | $\text{LogN}(1.434, 0.661)$ | [6] |
| $1/\eta$ | Asymptomatic Period | $\text{Gamma}(5, 1)$ | [7, 8] |
| $1/\theta$ | Presymptomatic Period | $\text{Gamma}(1.058, 2.174)$ | [9] |
| $1/\gamma$ | Infectious period | $\text{Gamma}(2.768, 1.156)$ | [7, 8] |
| $f_a$ | % of symptomatic cases who self-isolate | 80% | Assumed |
| $1/\tau$ | Average time to self-isolation post-symptom onset | 24 hours | Assumed |
| $\varepsilon_1, \varepsilon_2$ | Vaccine efficacy in preventing infection after first and second dose | 46%, 60% | [10, 11] |
| $\rho_{\{1,2\}}$ | Vaccine efficacy against severe symptomatic disease (1st and 2nd dose) | 66%, 94% | [10, 11] |

[1] calculated by subtracting the sampled $1/\theta$ from the sampled incubation period.
[*] The Gamma distributions are parametrized by shape and scale.

### 2 SUPPLEMENTARY RESULTS

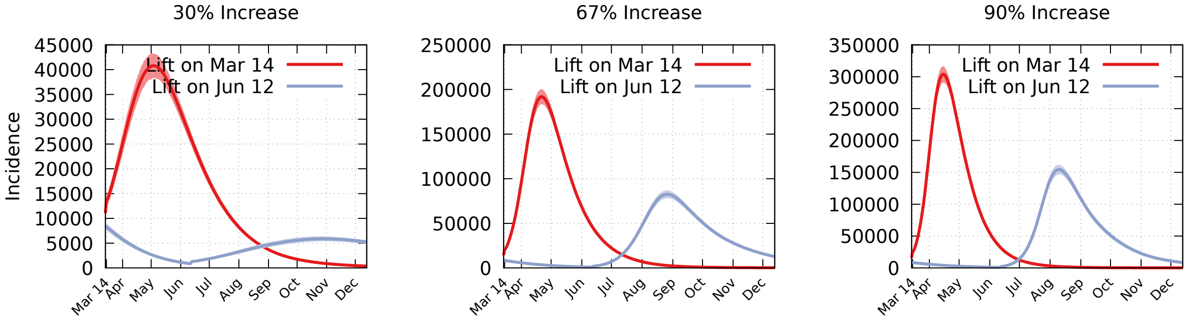


**(a)**

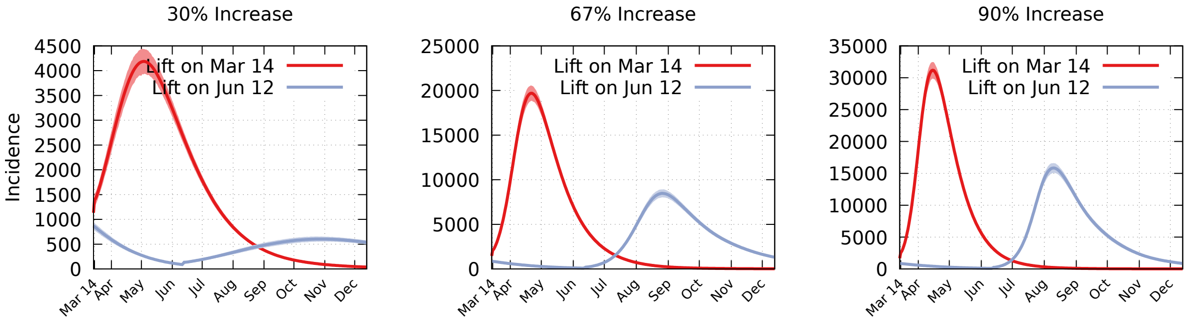


**(b)**

**Figure A3.** Projected incidence curves for (a) Texas and (b) Mississippi through the end of 2021. Each column denotes the increase in transmission starting on March 14.

**Table A3.** Cumulative infections, hospitalizations, and deaths expected for Texas between March 14 and December 31, 2021 under scenarios in which non-pharmaceutical interventions (NPI) are lifted on March 14 or June 12, and across variation in the degree to which transmission increases following that lift.

| $\beta$ increase by | March 14 | June 12 |
|---|---|---|
| | **Texas** | |
| | *Total Infections* | |
| 30% | 3,968,677 (3,791,011 - 4,136,566) | 1,125,259 (1,047,637 - 1,203,243) |
| 67% | 11,392,446 (11,220,054 - 11,553,711) | 7,344,552 (7,130,146 - 7,549,308) |
| 90% | 14,379,370 (14,246,099 - 14,508,837) | 10,824,185 (10,637,085 - 11,028,447) |
| | *Hospitalizations* | |
| 30% | 52,164 (49,719 - 54,665) | 10,987 (10,296 - 11,687) |
| 67% | 170,909 (167,454 - 174,379) | 60,122 (58,299 - 61,883) |
| 90% | 230,164 (227,066 - 233,176) | 97,378 (95,495 - 99,276) |
| | *Deaths* | |
| 30% | 1583 (1506 - 1668) | 409 (380 - 441) |
| 67% | 5647 (5511 - 5804) | 2145 (2071 - 2210) |
| 90% | 8231 (8058 - 8404) | 3514 (3435 - 3598) |
| | **Mississippi** | |
| | *Total Infections* | |
| 30% | 407,325 (389,090 - 424,556) | 115,491 (107,524 - 123,495) |
| 67% | 1,169,262 (1,151,569 - 1,185,813) | 753,807 (731,801 - 774,822) |
| 90% | 1,475,824 (1,462,146 - 1,489,112) | 1,110,939 (1,091,736 - 1,131,903) |
| | *Hospitalizations* | |
| 30% | 5354 (5103 - 5611) | 1128 (1057 - 1200) |
| 67% | 17,541 (17,187 - 17,897) | 6171 (5984 - 6351) |
| 90% | 23,623 (23,305 - 23,932) | 9994 (9801 - 10,189) |
| | *Deaths* | |
| 30% | 162 (155 - 171) | 42 (39 - 45) |
| 67% | 580 (566 - 596) | 220 (213 - 227) |
| 90% | 845 (827 - 863) | 361 (353 - 369) |
